# Large-Scale Evaluation of the Positive and Negative Syndrome Scale (PANSS) Symptom Architecture in Schizophrenia

**DOI:** 10.1101/2020.08.10.20170662

**Authors:** Keane Lim, Oon-Him Peh, Zixu Yang, Gurpreet Rekhi, Attilio Rapisarda, Yuen-Mei See, Nur Amirah Abdul Rashid, Mei-San Ang, Sara-Ann Lee, Kang Sim, Hailiang Huang, Todd Lencz, Jimmy Lee, Max Lam

**Affiliations:** Research Division, Institute of Mental Health, Singapore; Duke-NUS Medical School, Singapore; Stanley Center for Psychiatric Research, The Broad Institute of MIT and Harvard, Cambridge, Massachusetts; Feinstein Institute of Medical Research, The Zucker Hillside Hospital, New York; Department of Psychosis, Institute of Mental Health, Singapore; Neuroscience and Mental Health, Lee Kong Chian School of Medicine, Nanyang Technological University, Singapore

**Author notes:** Authors contributed equally to this work (Keane Lim, Oon-Him Peh). Authors co-supervised this work (Jimmy Lee, Max Lam). **Corresponding Author:** Max Lam, PhD Address: Research Division Institute of Mental Health, Singapore 10 Buangkok View Singapore 539747.

**Keywords:** Schizophrenia, PANSS, Factor structure, Exploratory factor analysis, Meta-analysis

## Abstract

Although the Positive and Negative Syndrome Scale (PANSS) is widely utilized in schizophrenia research, variability in specific item loading exist, hindering reproducibility and generalizability of findings across schizophrenia samples. We aim to establish a common metric PANSS factor structure from a large multi-ethnic sample and validate it against a meta-analysis of existing PANSS models. Schizophrenia participants (N = 3511) included in the current study were part of the Singapore Translational and Clinical Research Program (STCRP) and the Clinical Antipsychotic Trials for Intervention Effectiveness (CATIE). Exploratory Factor Analysis (EFA) was conducted to identify the factor structure of PANSS and validated with a meta-analysis (N = 16,171) of existing PANSS models. Temporal stability of the PANSS model and generalizability to individuals at ultra-high risk (UHR) of psychosis were evaluated. A five-factor solution best fit the PANSS data. These were the i) Positive, ii) Negative, iii) Cognitive/disorganization, iv) Depression/anxiety and v) Hostility factors. Convergence of PANSS symptom architecture between EFA model and meta-analysis was observed. Modest longitudinal reliability was observed. The schizophrenia derived PANSS factor model fit the UHR population, but not vice versa. We found that two other domains, Social Amotivation (SA) and Diminished Expression (DE), were nested within the negative symptoms factor. Here, we report one of the largest transethnic factorial structures of PANSS symptom domains (N = 19,682). Evidence reported here serves as crucial consolidation of a common metric PANSS that could aid in furthering our understanding of schizophrenia.

## Introduction

The Positive and Negative Syndrome Scale (PANSS) is a widely used 30-item clinician-rated instrument developed to provide comprehensive and reliable assessment of schizophrenia psychopathology.^1–5^ The PANSS was originally conceptualized to measure three domains of schizophrenia symptomatology (Positive, Negative and General psychopathology). Subsequent factor analytic reports indicated four-factor,^6^ five-factor,^7^ six-factor^8^ and seven-factor^9^ structures. The five-factor model consists of the Positive, Negative, Cognitive/disorganization, Depression/anxiety, and Hostility/excitement factors, is most commonly utilized for investigations on treatment response,^10^ functioning,^11^ insight,^12^ related psychotic disorders,^13^ cognition^14^ and social cognition.^15^

Reproducibility of specific item loading within the PANSS five-factor structure have been equivocal.^16,17^ Notable efforts were made to establish a consensus structure, to improve replicability and reproducibility of symptom measures.^17^ While the five-factor consensus model have been validated in Brazilian^18^ and Chinese^19,20^ samples separately, few reports have evaluated transethnic PANSS symptom architecture.^21^ Nevertheless, there has been much variability in the analytic approaches, dearth of independent sample validation, transethnic comparisons and sparse large scale reports that support the consensus factor structure of PANSS.^16,17^

Factor analytic studies in PANSS provide an avenue to establish measurable dimensional constructs in individuals with psychosis, which dovetails increasing efforts to study the dimensionality of psychopathology.^22^ Exploratory Factor Analysis (EFA) on PANSS items,^17,20^ facilitated symptom definition of psychopathology in schizophrenia. We had previously reported the possibility of further breaking down the definition of such symptom domains to evaluate subtler aspects of schizophrenia symptoms.^23,24^ The negative symptom factor is a candidate to examine. Negative symptomatology in schizophrenia appears to be a separable construct,^25–27^ relatively stable over time,^28^ and affects functioning more than positive symptoms.^29^ There is also evidence to suggest that negative symptoms could be further deconstructed into social amotivation and diminished expression. Broadly, the former implicates avolition for social activities and the latter expressive deficits.^30^ We demonstrated previously that components of negative symptoms were associated with cognition in a sizable number of schizophrenia individuals^23^ and were also likely to predict functioning outcomes in individuals at ultra-high risk of psychosis (UHR).^24^

The PANSS has been proven to be relatively sensitive to symptom changes, with a ‘past one week’ rating criteria.^1–4^ Nevertheless, it is likely that there are factors that tend to be more stable^31–34^ (e.g. negative and disorganized symptoms) while others are more amenable to changes over time. This warrants longer term prospective psychometric investigations of the PANSS factor structure.

### Aims and Hypothesis

The current study has three aims: (1) To establish evidence for a consensus PANSS factor structure and extend investigations to elucidate the two-factor structure within the negative symptoms factor. The underlying factor structure is obtained using EFA on our PANSS transethnic dataset, which is one of the largest reported. Based on earlier studies, we expect a five-factor PANSS structure that comprises of i) Positive, ii) Negative, iii) Cognitive/disorganization, iv) Depression/anxiety and v) Hostility factors. In order to comprehensively evaluate the factor structure, it is validated against a meta-analysis of existing PANSS models. (2) To determine the generalizability of PANSS factor structure to other related psychoses, particularly schizophrenia and UHR individuals. (3) To examine the longitudinal and configural stability of the factor structure. Here, we aim to examine the temporal stability of specific PANSS factors derived earlier in the study to further understand the phenomenological nature of psychosis.

## Methods

### Participants

Three cohorts were part of the Singapore Translational and Clinical Research in Psychosis program (STCRP) and the fourth cohort was from the Clinical Antipsychotic Trials for Intervention Effectiveness (CATIE) study. Details of the STCRP^20,23,24,35–38^ and the CATIE^39,40^ have been previously reported.

Two STCRP schizophrenia cohorts were of Asian descent and were recruited between 2005 to 2008 (N = H44)^20,36^ and 2008 to 2011 (N = 921).^23,35^ The third cohort were individuals (N = 168) deemed as Ultra-High Risk (UHR) of psychosis recruited between 2008 to 2011.^24,37,38,41^ Participants with history of neurological injuries, mental retardation and substance abuse were excluded. Diagnosis of schizophrenia was ascertained with the Structured Clinical Interview for DSM-IV-TR Axis I Disorder, Patients Edition.^42^ UHR individuals were ascertained via the Comprehensive Assessment of At-Risk Mental State (CAARMS).^43^ All studies were approved by the National Healthcare Group’s Domain Specific Review Board and written informed consent was obtained from all participants.

The fourth cohort was drawn from the CATIE study (N = 1446).^39,40^ The CATIE study was a NIMH funded multi-phase randomized controlled trial comparing the efficacy of typical and atypical antipsychotic medications, conducted between 2001 to 2004. Participants aged between 18 and 65 years were followed-up for up to 18 months. Exclusion criteria included diagnoses of schizoaffective disorder, first episode schizophrenia, mental retardation, and other cognitive disorders. The diagnosis of schizophrenia was ascertained via the Structured Clinical Interview for DSM-IV Axis I Disorder.^42^

### Statistical Analysis

#### Exploratory Factor Analysis (EFA): Deriving PANSS Factor Structure and Examining the Negative Symptoms Domain

EFA models (3 to 7 factor models) from combined STCRP and CATIE schizophrenia samples were extracted in Mplus 7.4. Listwise deletion was applied to participants with missing PANSS data. PANSS items were treated as ordinal variables and were subjected to both a cf-varimax and cf-equamax rotation with Weighted Least Squares Means and Variance adjusted estimation (WLSMV). Both varimax and equamax approaches were explored as they were the most commonly reported analysis approaches.^17^ To ensure that an unbiased data-driven approach was adopted, the adequacy of rotations (orthogonal or oblique) were assessed, by examining the inter-factor correlations on the combined sample.^44^ Factor correlation of r ≥ .32 would suggest that an oblique solution is appropriate.^44^ Model fit was assessed with multiple goodness-of-fit indices: Comparative Fit Index (CFI > .9), Tucker Lewis Index (TLI > .9), Root Mean Square Error of Approximation (RMSEA < .06), and Standardized Root Mean Square Residual (SRMR < .08).^45–47^ The Kaiser criterion^48^ of eigenvalue > 1 was also generated to aid in factor model extraction. Items with a factor loading ≥ .4 were assigned to the derived factor. Once the most parsimonious model was determined, the stability of the factor structure was evaluated by sequentially removing subsets of the combined schizophrenia sample, and repeating EFA iteratively.

To further examine the factor structure of negative symptoms, additional EFA was conducted using items with factor loading ≥ .4 on the negative symptoms factor. This was performed based on the factor structure derived from the most parsimonious model.

### Literature Review and Meta-analytic Procedure

A search of English-language studies was conducted in PubMed for studies published up to August 2018, along with combinations of the following search terms: “PANSS”, “five-factor”, “factor model”, “factor analysis”, “exploratory factor analysis”. The inclusion criteria were articles that reported the five-factor PANSS models in the schizophrenia spectrum (i.e., schizophrenia, schizoaffective, or schizophreniform) and available factor loadings ≥ .4. Articles that performed confirmatory factor analyses or were derived from overlapping samples were excluded. Factor loadings from these retrieved studies were extracted and meta-analysis of EFA findings were carried out.

The meta-analytic procedure involved extracting factor loadings from published reports. Thereafter, a co-occurrence matrix was computed by identifying the number of times a pair of PANSS items had their highest factor loadings on the same factor.^49,50^ Items with factor loading ≥ .4 were given a score of 1; otherwise a score of 0 was given. Secondary loadings and items with equal cross-loading were excluded. The co-occurrence for each pair of items was then standardized across studies by computing an index of similarity (Jaccard’s coefficient). This was calculated by dividing the number of times a pair of items had their highest loadings on the same factor by the total number of times the item was measured. The standardized cooccurrence matrix was then subjected to EFA with cf-varimax, unweighted least squares, oblique rotation in Mplus 7.4. This allows easy aggregation of published studies without additional raw data, correlation matrix or inter-factor correlation matrix. The approach is particularly suited for this context as factor loadings for all 30 PANSS items are often not reported in EFA reports.

### Comparison of Meta-analytic and Current Sample Factor Structure

Reliability and agreement of the PANSS factor structures derived from actual data and the literature review/meta-analysis were assessed with Pearson’s r and one-way random single measure intra-class correlation (ICC). Pearson’s r was computed by comparing the 30 factor loadings of each factor derived. The ICC of the data derived and literature/meta-analysis derived PANSS factor structure were investigated by computing both a summed average factor score and a weighted average factor score of items with factor loadings ≥ .4.^51^

### Generalizability of PANSS: Comparisons between Schizophrenia and UHR

To investigate the generalizability of the PANSS factors to related conditions of psychosis, we examined if PANSS factor scores derived from PANSS factor structures could differentiate schizophrenia and UHR status. Weighted average factor scores and summed average scores were computed for items with factor loadings ≥ .4. Weighted average factor scores were computed for each factor derived from the most parsimonious model by taking the average sum of the product of items with factor loadings ≥ .4 by the raw PANSS items score. We recently derived the PANSS factor structure for UHR individuals;^38^ the available factor scores were utilized in the current study. Logistic regression was then conducted on the schizophrenia-UHR status as the dependent variable and the PANSS derived factor scores as predictors. Hence, a total of 10 binary logistic models (5 factors × 2 group status) were performed for each factor score type (i.e., weighted vs summed average factor scores). A Bonferroni adjusted alpha level of .01 (.05/5) was applied for each factor score type. All analyses were conducted in IBM SPSS 20, unless otherwise stated. Confirmatory factor analysis (CFA) was also conducted to examine differential factor structures between UHR and schizophrenia. CFA was performed in Mplus 7.4.

### Longitudinal Reliability of PANSS Domains

Both weighted average factor scores and summed average factor scores were used to examine the stability of PANSS over time in participants with complete PANSS follow-up data. Spearman’s rank partial correlations were performed between the initial and follow-up PANSS factors scores, while controlling for the duration of follow-up period. Thereafter, these correlation coefficients were compared using Steiger’s z-test^52^ and Zou’s 95% confidence interval^53^ for non-overlapping correlations with dependent groups. The inclusion of zero in the 95% confidence interval would indicate no statistical differences between the two correlation coefficients of interest. This analysis was performed using the ‘cocor’ statistical program.^54^

### Data Availability

The data that support the findings of this study are available from the corresponding author upon reasonable request.

## Results

### Descriptive Statistics of Study Samples

The sample characteristics for schizophrenia samples are reported in Table 1. A total of N = 3511 individuals was included in the current EFA analysis. Demographic details of the UHR sample were as reported by Yang et al (2018).^38^

**Table 1.**
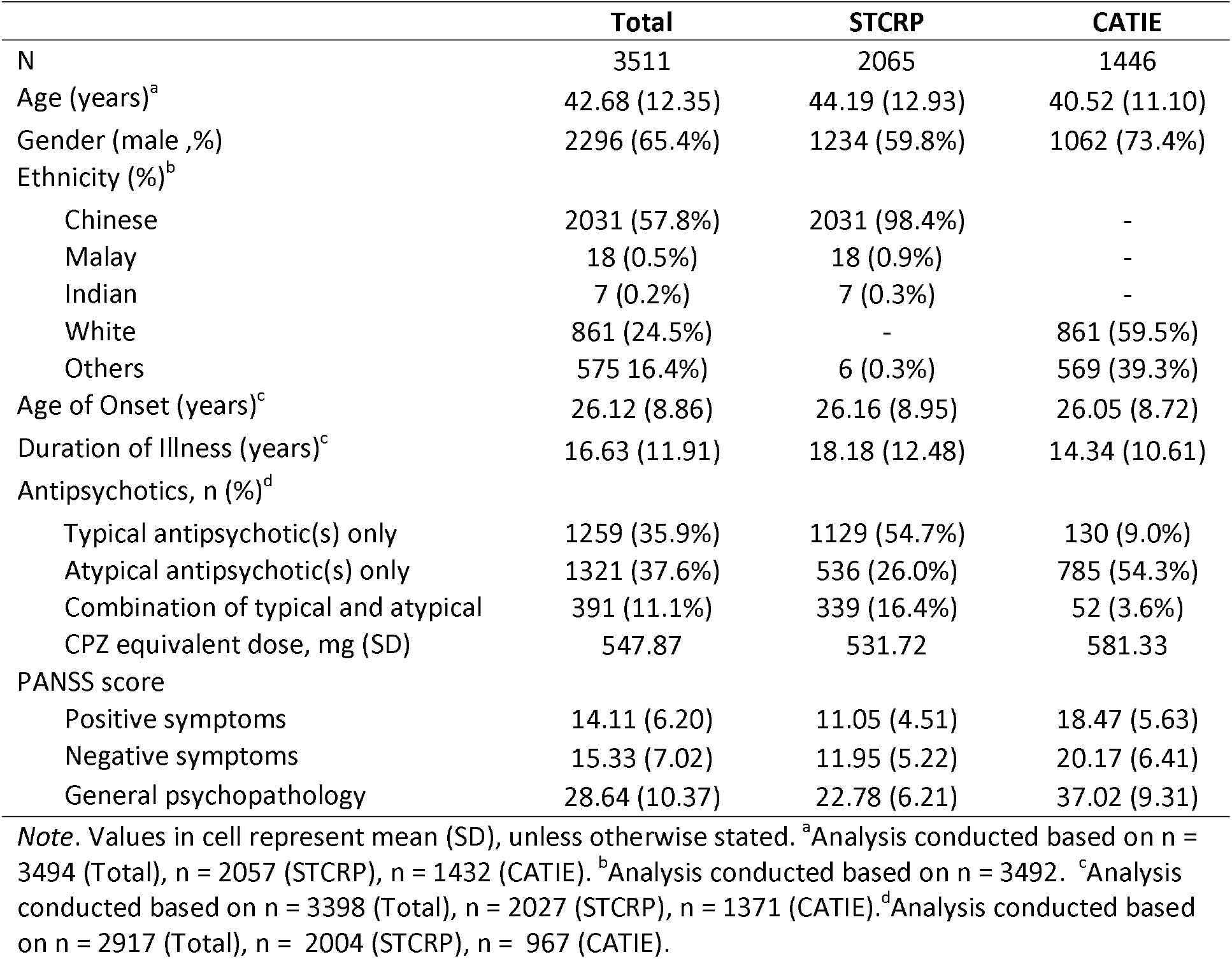
Sample demographics and clinical characteristics of schizophrenia individuals from STCRP and CATIE.

|  | Total | STCRP | CATIE |
| --- | --- | --- | --- |
| N | 3511 | 2065 | 1446 |
| Age (years)<sup>a</sup> | 42.68 (12.35) | 44.19 (12.93) | 40.52 (11.10) |
| Gender (male ,%) | 2296 (65.4%) | 1234 (59.8%) | 1062 (73.4%) |
| Ethnicity (%)<sup>b</sup> |  |  |  |
| Chinese | 2031 (57.8%) | 2031 (98.4%) | - |
| Malay | 18 (0.5%) | 18 (0.9%) | - |
| Indian | 7 (0.2%) | 7 (0.3%) | - |
| White | 861 (24.5%) | - | 861 (59.5%) |
| Others | 575 (16.4%) | 6 (0.3%) | 569 (39.3%) |
| Age of Onset (years)<sup>c</sup> | 26.12 (8.86) | 26.16 (8.95) | 26.05 (8.72) |
| Duration of Illness (years)<sup>c</sup> | 16.63 (11.91) | 18.18 (12.48) | 14.34 (10.61) |
| Antipsychotics, n (%)<sup>d</sup> |  |  |  |
| Typical antipsychotic(s) only | 1259 (35.9%) | 1129 (54.7%) | 130 (9.0%) |
| Atypical antipsychotic(s) only | 1321 (37.6%) | 536 (26.0%) | 785 (54.3%) |
| Combination of typical and atypical | 391 (11.1%) | 339 (16.4%) | 52 (3.6%) |
| CPZ equivalent dose, mg (SD) | 547.87 | 531.72 | 581.33 |
| PANSS score |  |  |  |
| Positive symptoms | 14.11 (6.20) | 11.05 (4.51) | 18.47 (5.63) |
| Negative symptoms | 15.33 (7.02) | 11.95 (5.22) | 20.17 (6.41) |
| General psychopathology | 28.64 (10.37) | 22.78 (6.21) | 37.02 (9.31) |

*Note.* Values in cell represent mean (SD), unless otherwise stated. <sup>a</sup>Analysis conducted based on n = 3494 (Total), n = 2057 (STCRP), n = 1432 (CATIE). <sup>b</sup>Analysis conducted based on n = 3492. <sup>c</sup>Analysis conducted based on n = 3398 (Total), n = 2027 (STCRP), n = 1371 (CATIE). <sup>d</sup>Analysis conducted based on n = 2917 (Total), n = 2004 (STCRP), n = 967 (CATIE).

### Exploratory Factor Analysis and Meta-analysis: PANSS Five Factor Architecture

Fit indices (Table 2) indicated that PANSS factor models with 4-7 domains of PANSS fit the overall schizophrenia sample data. Together with eigenvalues (> 1), the 5-factor model was selected. A trend where most inter-factor correlations exhibited r > .32 was found (Supplementary Table 1), this suggests that an oblique solution likely fit the data better than an orthogonal solution. Subsequently, a series of 5-factor models with oblique rotation, using cf-varimax and cf-equamax, were performed by sequentially removing subsets of the combined sample, to examine the change(s) in PANSS architecture. These results (Supplementary Tables 2-3) indicated that the cf-varimax solution provided a better fit, showing fewer cross-loadings when subsets of samples were removed. For comparison, orthogonal rotations are also presented in Supplementary Tables 4-5. Models with orthogonal rotation had more cross-loadings compared to an oblique solution, reducing interpretability of the PANSS factor structure. The optimal 5-factor model with cf-varimax oblique rotation is presented in Table 3. The five identified factors were (1) Positive, (2) Negative, (3) Cognitive/disorganization, (4) Depression/anxiety, and (5) Hostility.

**Table 2.** Comparison of model fit indices.

| Factor model | 3 | 4 | 5 | 6 | 7 |
| --- | --- | --- | --- | --- | --- |
| RMSEA | 0.073 | 0.057 | **0.048** | 0.042 | 0.037 |
| CFI | 0.938 | 0.965 | **0.977** | 0.984 | 0.989 |
| TLI | 0.922 | 0.952 | **0.967** | 0.974 | 0.98 |
| SRMR | 0.047 | 0.034 | **0.026** | 0.022 | 0.019 |
| Eigenvalue | 1.764 | 1.336 | **1.033** | 0.881 | 0.73 |

*Note.* Fit indices are exactly the same for cf-varimax and cf-equamax with oblique and orthogonal rotation. Taking together evidence from fit indices and eigenvalues, PANSS five-factor model (in bold) is considered the best fit.

**Table 3.** Factor loadings of optimal 5-factor PANSS model (cf-varimax, oblique)

|  | STCRP + CATIE (N = 3511) |  |  |  |  |
| --- | --- | --- | --- | --- | --- |
|  | Pos | Neg | Cog | Dep | Hos |
| **Positive factor** |  |  |  |  |  |
| P1 Delusions | **0.923** | 0.019 | -0.009 | 0.010 | 0.026 |
| G9 Unusual thought content | **0.740** | -0.029 | 0.239 | 0.005 | -0.002 |
| P6 Suspiciousness/persecution | **0.605** | 0.180 | -0.152 | 0.187 | 0.209 |
| P3 Hallucinatory behavior | **0.571** | 0.099 | 0.029 | 0.143 | 0.038 |
| P5 Grandiosity | **0.550** | -0.160 | 0.203 | 0.012 | 0.183 |
| **Negative factor** |  |  |  |  |  |
| N2 Emotional withdrawal | 0.195 | **0.721** | 0.011 | 0.159 | 0.057 |
| N4 Passive/apathetic social withdrawal | 0.196 | **0.713** | -0.043 | 0.203 | 0.068 |
| N6 Lack of spontaneity and flow of conversation | -0.159 | **0.709** | 0.282 | -0.071 | 0.192 |
| N3 Poor rapport | -0.100 | **0.660** | 0.186 | -0.094 | 0.365 |
| N1 Blunted affect | 0.024 | **0.645** | 0.227 | -0.003 | -0.119 |
| G16 Active social avoidance | 0.271 | **0.536** | -0.098 | 0.292 | 0.119 |
| G7 Motor retardation | -0.035 | **0.514** | 0.365 | 0.27 | -0.159 |
| **Cognitive/disorganization factor** |  |  |  |  |  |
| P2 Conceptual disorganization | 0.300 | -0.06 | **0.565** | 0.033 | 0.148 |
| G11 Poor attention | 0.049 | 0.109 | **0.538** | 0.187 | 0.197 |
| N7 Stereotyped thinking | 0.235 | 0.029 | **0.495** | 0.106 | 0.163 |
| N5 Difficulty in abstract thinking | 0.066 | 0.236 | **0.483** | 0.028 | 0.089 |
| G13 Disturbance of volition | 0.006 | 0.261 | **0.471** | 0.114 | 0.093 |
| G5 Mannerism & posturing | 0.050 | 0.169 | **0.465** | 0.271 | 0.019 |
| G15 Preoccupation | 0.217 | 0.070 | **0.460** | 0.165 | 0.174 |
| **Depression/anxiety factor** |  |  |  |  |  |
| G2 Anxiety | -0.001 | -0.073 | 0.048 | **0.835** | 0.077 |
| G3 Guilt feelings | 0.078 | 0.032 | 0.003 | **0.672** | 0.078 |
| G6 Depression | 0.077 | 0.165 | -0.039 | **0.662** | 0.044 |
| G4 Tension | -0.005 | 0.045 | 0.168 | **0.614** | 0.206 |
| G1 Somatic concern | 0.158 | -0.036 | 0.177 | **0.421** | 0.080 |
| **Hostility factor** |  |  |  |  |  |
| P7 Hostility | 0.073 | 0.018 | -0.105 | 0.135 | **0.813** |
| G8 Uncooperativeness | -0.032 | 0.154 | 0.114 | -0.049 | **0.713** |
| G14 Poor impulse control | 0.076 | -0.091 | 0.088 | 0.146 | **0.626** |
| P4 Excitement | 0.168 | -0.267 | 0.234 | 0.204 | **0.464** |
| **Items with loadings < 0.4** |  |  |  |  |  |
| G10 Disorientation | 0.005 | 0.184 | 0.373 | -0.025 | 0.103 |
| G12 Lack of judgment and insight | 0.266 | 0.117 | 0.324 | -0.289 | 0.175 |
| **Inter-factor correlations** |  |  |  |  |  |
| Pos | 1 |  |  |  |  |
| Neg | 0.162 | 1 |  |  |  |
| Cog | 0.307 | 0.409 | 1 |  |  |
| Dep | 0.477 | 0.308 | 0.269 | 1 |  |
| Hos | 0.434 | 0.307 | 0.442 | 0.448 | 1 |

*Note.* Pos = Positive factor, Neg = Negative factor, Cog = Cognitive/disorganization factor, Dep = Depression/anxiety factor, Hos = Hostility factor. Loadings $\geq 0.4$ are in bold.

The systematic search of PANSS factor analysis reports yielded 142 records. The title, abstract, and full text of these records were reviewed based on the inclusion criteria. The reference list of these papers was also reviewed. A total of 33 reports across 16,171 participants were included in this analysis^6,8,9,18,19,55–79^. Characteristics of the included studies are presented in Supplementary Table 6. The meta-analysis results and summary of item counts are presented in Table 4 and Supplementary Table 7 respectively. Meta-analytic results clearly indicated that a five-factor model is largely congruent with the EFA results within our samples.

**Table 4.** Five-factor solution of PANSS meta-analysis (N = 16,171)

|  | Pos | Neg | Cog | Dep | Hos |
| --- | --- | --- | --- | --- | --- |
| Positive factor |  |  |  |  |  |
| P1 Delusions | **0.982** | 0.000 | -0.027 | -0.002 | -0.029 |
| P3 Hallucinatory behavior | **0.921** | 0.001 | -0.03 | -0.007 | -0.001 |
| G9 Unusual thought content | **0.837** | -0.015 | 0.074 | -0.001 | -0.022 |
| P6 Suspiciousness/persecution | **0.756** | 0.005 | -0.016 | 0.004 | 0.040 |
| P5 Grandiosity | **0.756** | 0.000 | 0.008 | -0.003 | 0.040 |
| G12 Lack of judgment and insight | **0.416** | 0.058 | 0.180 | 0.051 | 0.095 |
| Negative factor |  |  |  |  |  |
| N4 Passive/apathetic social withdrawal | -0.003 | **0.972** | 0.003 | 0.025 | 0.019 |
| N2 Emotional withdrawal | -0.001 | **0.969** | 0.007 | 0.008 | -0.011 |
| N6 Lack of spontaneity and flow of conversation | -0.002 | **0.956** | 0.030 | -0.024 | -0.007 |
| N1 Blunted affect | 0.000 | **0.955** | 0.011 | -0.022 | -0.014 |
| N3 Poor rapport | 0.015 | **0.912** | 0.008 | -0.011 | 0.011 |
| G7 Motor retardation | -0.011 | **0.800** | 0.071 | 0.030 | 0.016 |
| G16 Active social avoidance | -0.001 | **0.571** | 0.021 | 0.113 | 0.075 |
| Cognitive/disorganization factor |  |  |  |  |  |
| G11 Poor attention | -0.019 | -0.027 | **0.851** | -0.008 | -0.008 |
| P2 Conceptual disorganization | 0.019 | -0.048 | **0.844** | -0.009 | -0.022 |
| N5 Difficulty in abstract thinking | -0.023 | 0.047 | **0.810** | -0.016 | -0.029 |
| G10 Disorientation | 0.015 | -0.020 | **0.573** | 0.004 | -0.005 |
| N7 Stereotyped thinking | 0.027 | -0.023 | **0.535** | 0.014 | 0.122 |
| G13 Disturbance of volition | -0.007 | 0.250 | **0.486** | 0.020 | 0.006 |
| G5 Mannerism & posturing | 0.016 | 0.015 | **0.403** | -0.005 | 0.142 |
| Depression/anxiety factor |  |  |  |  |  |
| G6 Depression | -0.007 | 0.000 | -0.007 | **0.960** | -0.047 |
| G3 Guilt feelings | -0.006 | -0.008 | -0.004 | **0.934** | -0.045 |
| G2 Anxiety | 0.001 | 0.003 | -0.011 | **0.900** | 0.053 |
| Hostility factor |  |  |  |  |  |
| P7 Hostility | -0.002 | -0.014 | -0.014 | -0.010 | **1.000** |
| P4 Excitement | 0.001 | 0.015 | -0.024 | -0.006 | **0.950** |
| G14 Poor impulse control | -0.002 | -0.027 | 0.044 | -0.013 | **0.895** |
| G8 Uncooperativeness | 0.001 | 0.050 | -0.013 | -0.011 | **0.753** |
| G4 Tension | 0.000 | -0.027 | 0.056 | 0.414 | **0.418** |
| Items with loadings < 0.4 |  |  |  |  |  |
| G1 Somatic concern | 0.092 | -0.015 | 0.047 | 0.356 | 0.024 |
| G15 Preoccupation | 0.071 | 0.080 | 0.331 | 0.133 | 0.026 |

*Note.* Pos = Positive factor, Neg = Negative factor, Cog = Cognitive/disorganization factor, Dep = Depression/anxiety factor, Hos = Hostility factor. Loadings $\geq 0.4$ are in bold.

Pearson’s r comparing factor loadings of the PANSS factor structure derived from actual data and those from the meta-analysis were highly consistent (Positive: .902, *P* < .001; Negative: r = .927, *P* < .001; Cognitive/disorganization: r = .815, *P* < .001; Depression/anxiety: r = .850, *P* < .001; Hostility: r = .859, *P<* .001). Summed and weighted factor scores were derived from the both data derived and meta-analysis PANSS factor structure (See Methods). ICC analysis indicated greater agreement when average summed factor score was used compared to weighted average factor scores (Positive: ICC_sum_ = .966, ICC_weighted_ = −942; Negative: ICC_sum_ = 1, ICC_Weighted_ = −783; Cognitive/disorganization: ICC_sum_= .974, ICC_weighted_ = −771; Depression/anxiety: ICC_sum_ = −934, ICC_weighted_ = −622; Hostility: ICC_sum_= .951, ICC_weighted_ = −822; where all *P<* .001).

### Exploratory Factor Analysis: PANSS Negative Symptom Sub-Architecture

To investigate if the negative symptoms factor could be deconstructed further, a cf-varimax oblique rotation was performed using the negative symptoms items derived from the combined dataset. A two-factor solution was obtained (Supplementary Table 8): (1) Diminished Expression (DE), and (2) Social Amotivation (SA). The fit statistics indicated a good fit, CFI = .994, TLI = .983, RMSEA = .093, SRMR = .022. The DE factor comprised of N1 Blunted affect, N3 Poor rapport, N6 Lack of spontaneity and flow of conversation, and G7 Motor retardation. The SA factor comprised of N2 Emotional withdrawal, N4 Passive/apathetic social withdrawal, and G16 Active social avoidance. The inter-factor correlation was r = .644.

### Generalizability of PANSS Factor Structure on Schizophrenia-UHR Status

Results from logistic regression using factor scores derived from the current schizophrenia sample and the UHR factor structure^38^ are reported in Supplementary Table 9. Bonferroni corrected univariate logistic regression showed significant difference between schizophrenia and UHR models, except for the depression/anxiety factor.

Two CFA models were conducted to explore if the schizophrenia factor structure fit the UHR model and vice versa. The schizophrenia factor structure showed modest fit on the UHR population (RMSEA = .062, CFI = .890, HI = .878), but the UHR factor structure^38^ showed poor fit when applied on the schizophrenia sample (RMSEA = .105, CFI = .870, HI = .854).

### Longitudinal Reliability of PANSS

Prospective data was available for N = 1098 participants. The follow-up duration was 2 years for STCRP cohort (n = 116) and 6 months for CATIE cohort (n = 982). Relative stability of the PANSS factors showed strongest correlation observed for the Cognitive/disorganization factor (Supplementary Table 9 and Supplementary Figures 1-3). All associations remained significant after adjusting for Bonferroni correction (*P* <.01). Using the Cognitive/disorganization factor as the reference factor, time-based correlation coefficients were compared across factors. There were no significant differences between Cognitive/disorganization and Negative factors (Supplementary Table 10), and significant differences for Positive, Depression/anxiety and Hostility factors after Bonferroni correction (*P* < .0125). Further post-hoc analysis of the negative symptoms’ two factors indicated no difference between cognitive/disorganization and DE, and significant difference for SA, after Bonferroni correction (*P* < .025). Similar pattern of results was obtained for both weighted and summed average factor scores (Supplementary Table 10).

## Discussion

The current study examines comprehensively the PANSS factor structure in the largest consolidated transethnic sample (N = 19,682) in the literature. We investigated the derivation of symptom architecture, confirmed by a review and meta-analysis of existing studies, existence of sub-architecture within the PANSS, generalizability of the PANSS symptom architecture to a limited cohort of UHR individuals and temporal stability of the factor structure.

### Identifying PANSS Factor Structure via EFA and Meta-analysis

We identified five PANSS symptom domains: (1) Positive, (2) Negative, (3) Cognitive/disorganization, (4) Depression/anxiety, and (5) Hostility factors. The five-factor structure was robust within the cf-varimax oblique rotation framework, which optimized factor rotation by preventing factor collapse where inter-factor correlations tend to one;^80^ thus allowing the variance of PANSS items’ factor loadings to be maximized across the items and factors, providing a satisfactory interpretable solution. This is apparent when each dataset was left out iteratively and factor loadings appeared invariant even with smaller sample sizes. Unlike some earlier reports that generally assumed orthogonal structure within the PANSS,^17,56,68^ both orthogonal and oblique factor solutions were examined, the latter demonstrating superior fit. This is not a trivial finding. The oblique rotation provides more refined structure that incorporates inter-factor correlations^44^ which has higher ecological value and better represent the complex dimensional nature of symptoms in schizophrenia. Given the backdrop of high comorbidity^81^ and widespread biological associations^82^ reported between schizophrenia and other psychiatric conditions or traits, our efforts further support downstream research in these areas.

It was noted that factor loadings between EFA of actual data and those of meta-analysis were highly congruent. As such, we went a step further to compute factor scores using both models for individuals with PANSS data, using (i) unweighted and (ii) weighted approaches. There was better agreement in models for summed average factor score, compared to a weighted average factor score. It possible that summed average factor score allows easy generalizability across studies, as all items loading at a given cutoff within the factor were computed with an equal weighting scheme; the weighted factor score accounts implicitly for the strength (or lack of strength) of each item depending on loading magnitude, allowing data specific details that could prove useful in detecting precise differences for downstream association analysis.^51,83^ Nonetheless, subtle variability across and within studies may also be captured given the more sensitive nature of computing factor scores by weights.

### Generalizability of PANSS Factor Structure on Schizophrenia-UHR status

Factor scores derived from a UHR-specific^38^ and schizophrenia-specific PANSS factor structure largely differentiated schizophrenia-UHR status, with the exception of the depression/anxiety factor. Higher prevalence of comorbid affective disorders in the UHR population undifferentiated from individuals with psychosis had been previously reported by our group and others.^41,84^ CFA revealed that the schizophrenia model could be generalized to the UHR population but not vice versa. These results partially supports the idea that cooccurrence of non-specific, subthreshold psychotic symptoms and, non-psychotic symptoms are likely present in UHR samples^85,86^ but subtle phenomenological difference still exists when compared to schizophrenia. Alternatively, symptom structure derived from a much smaller sample might not be generalizable to the larger schizophrenia sample reported in the current study, while the vice versa might have been possible. Nonetheless, further research is necessary to clarify the suitability of applying PANSS factor structure derived in schizophrenia specific populations to other related psychoses.

### Longitudinal Reliability of PANSS

The longitudinal reliability analysis indicated modest temporal stability of the symptom structure and factor scores across time. This may not necessarily indicate a psychometric flaw in PANSS; but reflects the time specificity inherent to the reference period of PANSS administration. Amongst the derived factors, the cognitive/disorganization factor showed the highest stability that is least amenable to change over time.^87^ The current report is also amongst the first to demonstrate the relative long-term stability of the PANSS cognitive/disorganization, negative and diminished expression symptom factors in a large-scale schizophrenia study. Our data justifies further deconstructing negative symptoms into the two domains of SA and DE^88^ - the latter representing a unique construct with stronger association to neuropsychological function^23^, and the former indexing more of a social motivation construct.^89^ Given the lack of effective treatments for negative symptoms, the deconvolution and refinement of these two domains could provide opportunities for exploring neurobiological underpinnings and treatment options for schizophrenia symptoms.^90^

### Limitations

A limitation of the meta-analysis approach included schizophrenia samples of differing chronicity. Further, studies included in the meta-analysis were narrowed to reports which provided factor loadings. This is further limited by the lack of studies reporting full factor loadings and inter-factor correlation matrix precluding technical reproduction of the correlation matrix which could then be pooled to give both configural and metric invariance - which could help with improving the robustness of factor models.^91–93^ While EFA factors derived from schizophrenia data is largely the same as those in the meta-analysis findings, differential loadings of two items G12 Lack of Judgement and Insight and G4 Tension were observed. Notably, these items are close to the factor item definition cutoff of < .40 and are likely to be within range of noise in the data. These might benefit from more refined and sensitive approaches to clarify more precisely item level loadings on factors. Nevertheless, evidence between factor analytic models of the PANSS in a large schizophrenia sample, and those carried out in literature meta-analysis converged.

## Conclusions

In conclusion, the current report contributes to existing evidence by elucidating a robust five-factor PANSS model that confirms expert consensus via large-scale data. Symptom level deconstruction and establishing latent dimensions of schizophrenia symptoms, are crucial and in line with the dimensional initiatives.^22,94–96^ We report robust evidence for the existence of dual negative symptom dimensions within the PANSS and demonstrate potential phenomenological separation between UHR and schizophrenia. Evidence reported here serves as a crucial consolidation of a common metric PANSS that could aid in furthering our understanding of schizophrenia.

## Data Availability

All data related to the manuscript has been made available within the manuscript and/or supplementary materials.

## Acknowledgements

This research was supported by grants from the Ministry of Health Singapore, National Medical Research Council (Grant No.: NMRC/TCR/003/2008, NMRC/CG/004/2013). ML is supported by the National Medical Research Council Research Training Fellowship (Grant No.: MH095:003/008-1014). The authors thank the NIH for providing limited access datasets for the NIMH CATIE (ClinicalTrials.gov identifier NCT00014001, NIMH contract #N01MH90001).

## Contributors

JL and ML designed the study. KL and OHP conducted the literature review, data analyses, and wrote the first draft of the manuscript. All authors gave substantial comments, edited the manuscript and approved the final manuscript.

